# Multiplex Antibody Analysis of IgM, IgA and IgG to SARS-CoV-2 in Saliva and Serum from Infected Children and their Close Contacts

**DOI:** 10.1101/2021.03.22.21254120

**Authors:** Carlota Dobaño, Selena Alonso, Marta Vidal, Alfons Jiménez, Rocío Rubio, Rebeca Santano, Diana Barrios, Gemma Pons Tomas, María Melé Casas, María Hernández García, Mònica Girona-Alarcón, Laura Puyol, Natalia Rodrigo Melero, Carlo Carolis, Aleix Garcia-Miquel, Elisenda Bonet-Carne, Joana Claverol, Marta Cubells, Claudia Fortuny, Victoria Fumadó, Anna Codina, Quique Bassat, Carmen Muñoz-Almagro, Mariona Fernández de Sevilla, Eduard Gratacós, Luis Izquierdo, Juan José García-García, Ruth Aguilar, Iolanda Jordan, Gemma Moncunill

## Abstract

COVID-19 affects children to a lesser extent than adults but they can still get infected and transmit SARS-CoV-2 to their contacts. Field deployable non-invasive sensitive diagnostic techniques are needed to evaluate the infectivity dynamics of the coronavirus in pediatric populations and guide public health interventions.

We evaluated the utility of high-throughput Luminex-based assays applied to saliva samples to quantify IgM, IgA and IgG antibodies against five SARS-CoV-2 spike (S) and nucleocapsid (N) antigens in the context of a contacts and infectivity longitudinal study. We compared the antibody levels obtained in saliva versus serum/plasma samples from a group of children and adults tested weekly by RT-PCR over 35 days and diagnosed as positive (n=58), and a group of children and adults who consistently tested negative over the follow up period (n=61), in the Summer of 2020 in Barcelona, Spain.

Antibody levels in saliva samples from individuals with confirmed RT-PCR diagnosis of SARS-CoV-2 infection were significantly higher than in negative individuals and correlated with those measured in sera/plasmas. Higher levels of anti-S IgG were found in asymptomatic individuals that could indicate protection against disease in infected individuals. Higher anti-S IgG and IgM levels in serum/plasma and saliva, respectively, in infected children compared to infected adults could also be related to stronger clinical immunity in them. Among infected children, males had higher levels of saliva IgG to N and RBD than females. Despite overall correlation, individual clustering analysis suggested that responses that may not be detected in blood could be patent in saliva, and vice versa, and therefore that both measurements are complementary.

In addition to serum/plasma, measurement of SARS-CoV-2-specific saliva antibodies should be considered as a complementary non-invasive assay to better estimate the percentage of individuals who have experienced coronavirus infection. Saliva antibody detection could allow determining COVID-19 prevalence in pediatric populations, alternative to bleeding or nasal swab, and serological diagnosis following vaccination.

## INTRODUCTION

Since the start of the COVID-19 pandemic, it has become apparent that pediatric populations are less affected than adult or older populations^1^. Clinical presentation of SARS-CoV-2 infection is milder in children, with more proportion of asymptomatic cases^1^. One hypothesis for this lower severity of COVID-19 is the protective effect that antibodies from human coronaviruses of the common cold (HCoV), which are more prevalent in children, could exert on SARS-CoV-2 control, considering cross-reactivity between them^2,3^. Children could also be harboring lower viral loads, in part due to the lower expression of ACE2 receptor of the coronavirus^4^. This implies that highly sensitive techniques might be required for its accurate diagnosis.

In this context, a question of high interest has been whether children who become infected might be less efficient transmitters to their immediate contacts, as this has important implications for the management of outbreaks in schools, extracurricular activities, and holiday camps. Having field deployable diagnostic tools to monitor infectivity dynamics in school-like environments is therefore very relevant from the public health perspective.

The presence of the virus in nasal or nasopharyngeal samples can be detected with real time polymerase chain reaction (RT-PCR) and antigen-based sensitive methods, while prior exposure has to be assessed by detecting antibodies to the SARS-CoV-2 antigens. The latter is usually done by measuring specific immunoglobulins in serum or plasma samples, which requires obtaining blood samples by venous or capillary punctures, less amenable to large field studies. Several commercial rapid diagnostic tests (RDT) exist that are useful as point of care kits, but they may be limited by their sensitivity and specificity in the case of asymptomatic infections, which usually induce lower levels of antibodies^5,6^. A number of laboratory-based serological assays (ELISA, CLIA) are also widely used and with good performances. Similarly, multiplex assays combining multiple antigenic specificities simultaneously and amenable for high-throughput testing, offer even better potential to have the highest sensitivity to detect low-level responses in younger populations.

Being a respiratory pathogen, the mucosal immunity has a key importance, and thus the role of IgA in controlling the virus is becoming increasingly important^7^. Therefore, saliva is an attractive sample matrix for developing field-deployable non-invasive serological assays that are readily applicable in pediatric surveys. Indeed, antibodies in saliva have been detected in COVID-19 patients and correlate with plasma antibodies^8^. Consequently, the availability of highly sensitive and specific antibody assays for immunological profiling is valuable both for immune-epidemiological surveys and to better understand protective immunity to SARS-CoV-2. In addition, saliva serology would allow knowing the serological diagnosis after the vaccination for determining the levels of immunity at individual level and in the population, and could be really helpful in population screening for determining SARS-CoV-2 seroprevalence in children populations.

In this study we adapted and evaluated three Luminex-based antibody assays to quantify the levels of IgM, IgA and IgG against several SARS-CoV-2 antigens in saliva samples, and compared them to the levels of antibodies obtained using serum/plasma samples from the same individuals who had a positive or negative RT-PCR diagnosis over a five week follow up period. We tested the applicability of the Luminex saliva assays in children and adult volunteers, having different demographic characteristics and clinical presentation, participating in a contacts and infectivity study after the first peak of the COVID-19 pandemic in the Summer of 2020 in Barcelona.

## MATERIALS AND METHODS

### Study design, human subjects and samples

We compared the levels of SARS-CoV-2 antibodies in subjects with a positive diagnosis by nasopharyngeal RT-PCR and/or ELISA SARS-CoV-2 IgG, IgM test (Euroimmune Architect – Abbott) independently of the COVID-19 compatible symptoms or not, and subjects with negative nasopharyngeal RT-PCR^9^ and serology by RDT (SureScreen). Eligible subjects entered the study via three recruitment pathways: (i) active surveillance in 22 Summer schools, (ii) passive detection of cases coming from other school-like environments, referred from the Catalonian health surveillance system call, and (iii) individual cases referred from an announcement made to enroll children with positive RT-PCR in the previous 5 days. Participants were followed up for 5 weeks over July 2020, with weekly sample collection and RT-PCR testing that allowed accurately defining the infected positive and negative groups. Saliva samples were collected with Oracol devices (Malvern Medical Development, UK) for optimal harvesting of crevicular fluid, enriched with serum antibodies^10,11^. Blood samples were collected by venipuncture and plasma or serum separated by centrifugation and frozen, or as dried blood spots (DBS) to facilitate the field survey logistics.

Oracol devices were kept and transported refrigerated to the lab on the same day for centrifugation into a cryotube, heat inactivated for 30 min at 55°C, and frozen at -20°C until serological analysis. Thirty-six pre-pandemic plasmas from healthy adults were used as negative controls and to calculate the seropositivity cutoff for sera/plasmas. No pre-pandemic saliva samples were available after contacting national biobanks.

Study protocols for human subject research were approved by the Institutional Review Board and the Hospital Sant Joan de Déu Ethics Committee (Ref. PIC 153-20) and the Hospital Clinic ethics committee (Ref. CEIC-7455), and written informed consent was obtained from participants or guardians before starting study procedures.

### Measurement of antibodies

Quantitative suspension array technology (qSAT) assays to measure IgM, IgA and IgG against SARS-CoV-2 were adapted from our previous standardized serum/plasma protocols^12^ to a saliva matrix for SARS-CoV-2 antibody evaluation. Antigens included the nucleocapsid (N) full-length (FL) and C-terminus (amino acid residues 340-416, CT)^13^, the spike (S) FL produced at CRG, S2 purchased from SinoBiologicals, and RBD donated by F. Krammer (Mount Sinai, NY). Briefly, proteins coupled to magnetic microspheres (Luminex Corporation, Austin, TX) were incubated with serum/plasma (1/500 dilution) or saliva samples (1/5 or 1/10 dilutions, **Figure S1**) or blank controls in 384-well plates. Saliva dilution of 1/10 showed higher sensitivity and was selected for further assays. The impact of heat inactivation was previously checked in serum/plasma and saliva (**Figure S2**), with a decrease in the levels of IgM to RBD and S in saliva. Serum was eluted from DBS with 200 μl of PBS-BN (filtrated PBS with 1% BSA and 0.05% sodium azide, MilliporeSigma, St. Louis, USA) + 0.05% Tween20. Considering a hematocrit of 50% results in an eluted protein concentration equivalent to a serum/plasma dilution of 1:50, which was subsequently diluted to 1:500 for the assay. Antibodies in serum eluted from DBS and from serum/plasma samples are shown together since no differences were observed in the available paired samples of plasma and DBS. After antigen-coupled beads were incubated with samples, plates were washed and phycoerythrin-labeled secondary antibodies (anti-human IgG, IgM, or IgA, Moss) added. Finally, beads were washed, resupended and acquired in a FlexMap 3D xMAP® instrument. Crude median fluorescent intensities (MFI) and background fluorescence from blank wells were exported using the xPONENT software.

### Data analysis

Non-parametric Mann-Whitney U tests were used in boxplots to compare levels (log_10_MFI) of each antibody/antigen pair between study groups. Radar charts were used to compare median log_10_MFI of all antibodies together between study groups by Mann-Whitney U test. Heatmaps with hierarchical clustering (Euclidean method) were used to evaluate patterns of responses at the individual level depending on clinical and demographic variables. Due to the unavailability of pre-pandemic saliva samples, we explored calculating seropositivity cutoffs by the mean plus 3 standard deviations (SD) of pandemic negative samples for serum/plasma and for saliva samples (**Figure S3**). All analyses were performed at 5% significance level with R software version 4.0.2. The ggplot2 package was used to perform boxplot graphs^14^.

## RESULTS

The basic demographic characteristics of SARS-CoV-2 infected (n=58) and non-infected (n=61) individuals in whom saliva and serum/plasma samples were analyzed, are shown in **Table 1**.

**Table 1.** Characteristics of study participants.

|  | **Negatives** |  | **Positives** |  |
| --- | --- | --- | --- | --- |
|  | Serum <sup>a</sup><br>(N=48) | Saliva<br>(N=61) | Serum <sup>b</sup><br>(N=58) | Saliva<br>(N=56) |
| Age |  |  |  |  |
| Children | 5 (10.4%) | 10 (16.4%) | 40 (69.0%) | 40 (71.4%) |
| Adults | 43 (89.6%) | 51 (83.6%) | 18 (31.0%) | 16 (28.6%) |
| Sex |  |  |  |  |
| Male | 15 (31.2%) | 19 (31.1%) | 30 (51.7%) | 28 (50.0%) |
| Female | 33 (68.8%) | 42 (68.9%) | 28 (48.3%) | 28 (50.0%) |
| Symptoms |  |  |  |  |
| Yes | 2 (4.2%) | 2 (3.3%) | 26 (44.8%) | 26 (46.4%) |
| No | 46 (95.8%) | 59 (96.7%) | 32 (55.2%) | 30 (53.6%) |
| Sample collection (weeks) |  |  |  |  |
| 1 | 4 (20.0%) | 9 (34.6%) | 17 (32.1%) | 19 (34.5%) |
| 2 | 13 (65.0%) | 13 (50.0%) | 12 (22.6%) | 12 (21.8%) |
| 3 | 3 (15.0%) | 4 (15.4%) | 17 (32.1%) | 17 (30.9%) |
| 4 | 0 (0.0%) | 0 (0.0%) | 6 (11.3%) | 6 (10.9%) |
| 5 | 0 (0.0%) | 0 (0.0%) | 1 (1.9%) | 1 (1.8%) |

Adult: age 15 years or older
<sup>a</sup> Sixteen serum samples were obtained from dried blood spots (DBS)
<sup>b</sup> Eight serum samples were obtained from DBS

### Antibody levels according to SARS-CoV-2 RT-PCR status

We compared the levels (log_10_MFI) of IgM, IgA and IgG antibodies to five SARS-CoV-2 antigens in saliva and serum/plasma samples from RT-PCR positive and negative individuals. IgG levels to all antigens in saliva were statistically significantly higher in RT-PCR positive than negative individuals (**Figure 1A & 1B**). Levels of IgM and IgA to S (FL, S2 and RBD) but not N (FL, CT) antigens in saliva were statistically significantly higher in RT-PCR positive than negative individuals. In serum/plasma samples from the same individuals, all Ig isotypes were significantly higher in positive than negative individuals. The magnitude of antibody responses was substantially lower in saliva than serum/plasma samples despite being measured at a higher concentration (1/10 vs 1/500), and levels overlapped between positive and negative individuals to a higher degree in saliva than serum/plasma samples (**Figure S3**).

**Figure 1.**
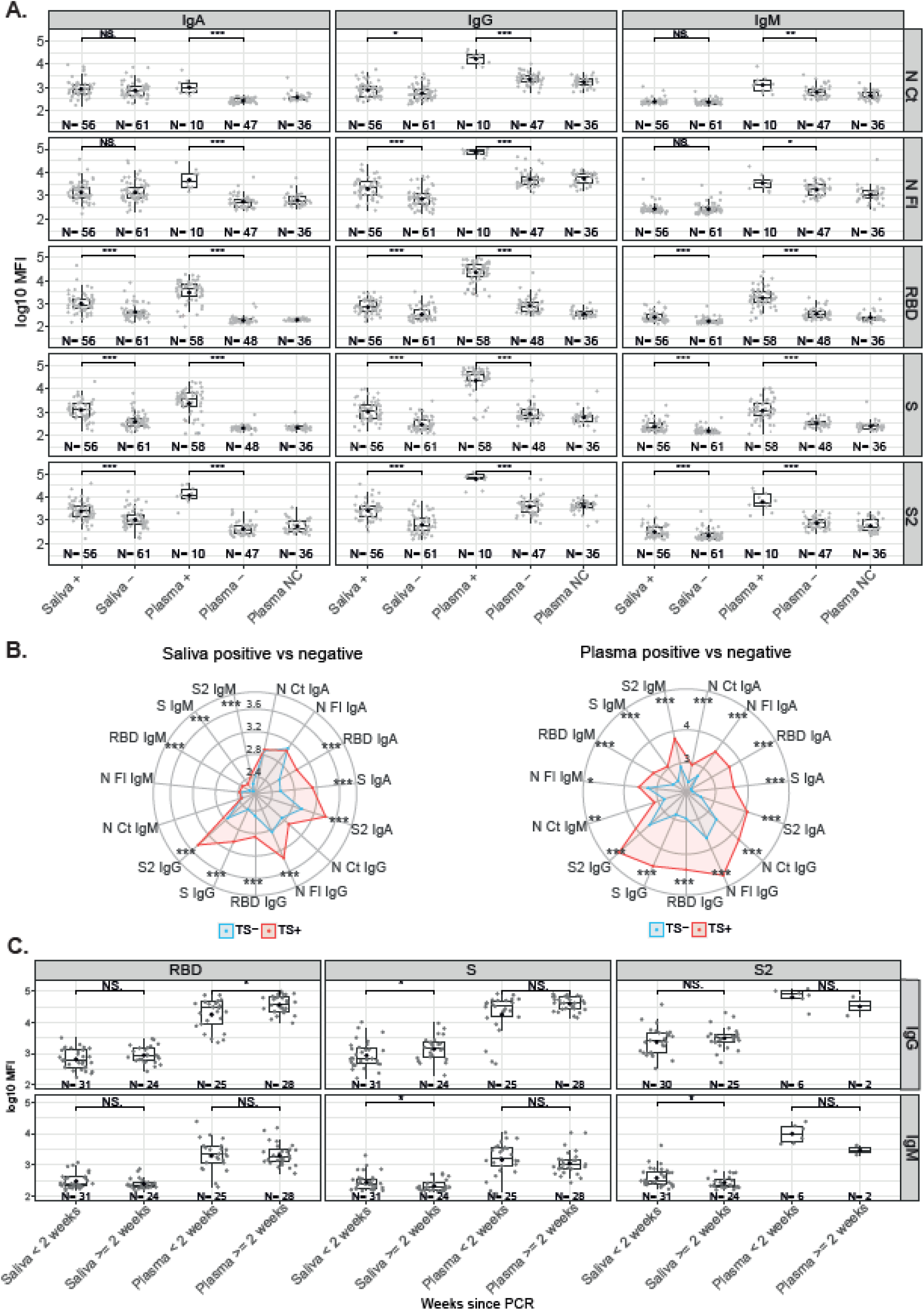
Antibody levels according to SARS-CoV-2 RT-PCR status. **A**. Boxplot showing log_10_MFI antibody levels. Saliva samples were tested heat inactivated and at 1/10 dilution, and serum (from plasma samples or dry blood spots) at 1/500. **B**. Radar charts representing the median of the log_10_MFI antibodies in plasma and saliva. TS-: Negative Test Sample, represented in blue. TS+: Positive Test Sample, represented in red. **C**. Boxplots showing log_10_MFI antibody levels by time since positive RT-PCR. Median log_10_MFI levels were compared by Mann-Whitney U test. Statistically significant raw p-values are highlighted with an asterisk. *** p<0.001, ** p<0.01, * p<0.05.

We evaluated whether RT-PCR positive individuals who were antibody negative had more recent infections. Stratified by time since diagnosis, levels of IgG against S in saliva and against RBD in serum/plasma were higher in samples collected >2 weeks after positive RT-PCR (**Figure 1C**). In contrast, levels of IgM to S and S2 in saliva were lower in samples collected >2 weeks after positive RT-PCR.

### Antibody levels by age and sex according to SARS-CoV-2 RT-PCR status

Among infected individuals, children had significantly higher serum/plasma levels of IgG to RBD and S, and significantly higher saliva levels of IgM to RBD and S than adults (**Figure 2**). In contrast, infected adults had significantly higher levels IgA to N FL in saliva than infected children. Infected male children also had higher saliva levels of IgG to N CT, N FL and RBD than female children (**Figure S4**). Among SARS-CoV-2 RT-PCR negative individuals, children compared to adults had significantly higher serum/plasma levels of IgG to N FL.

**Figure 2.**
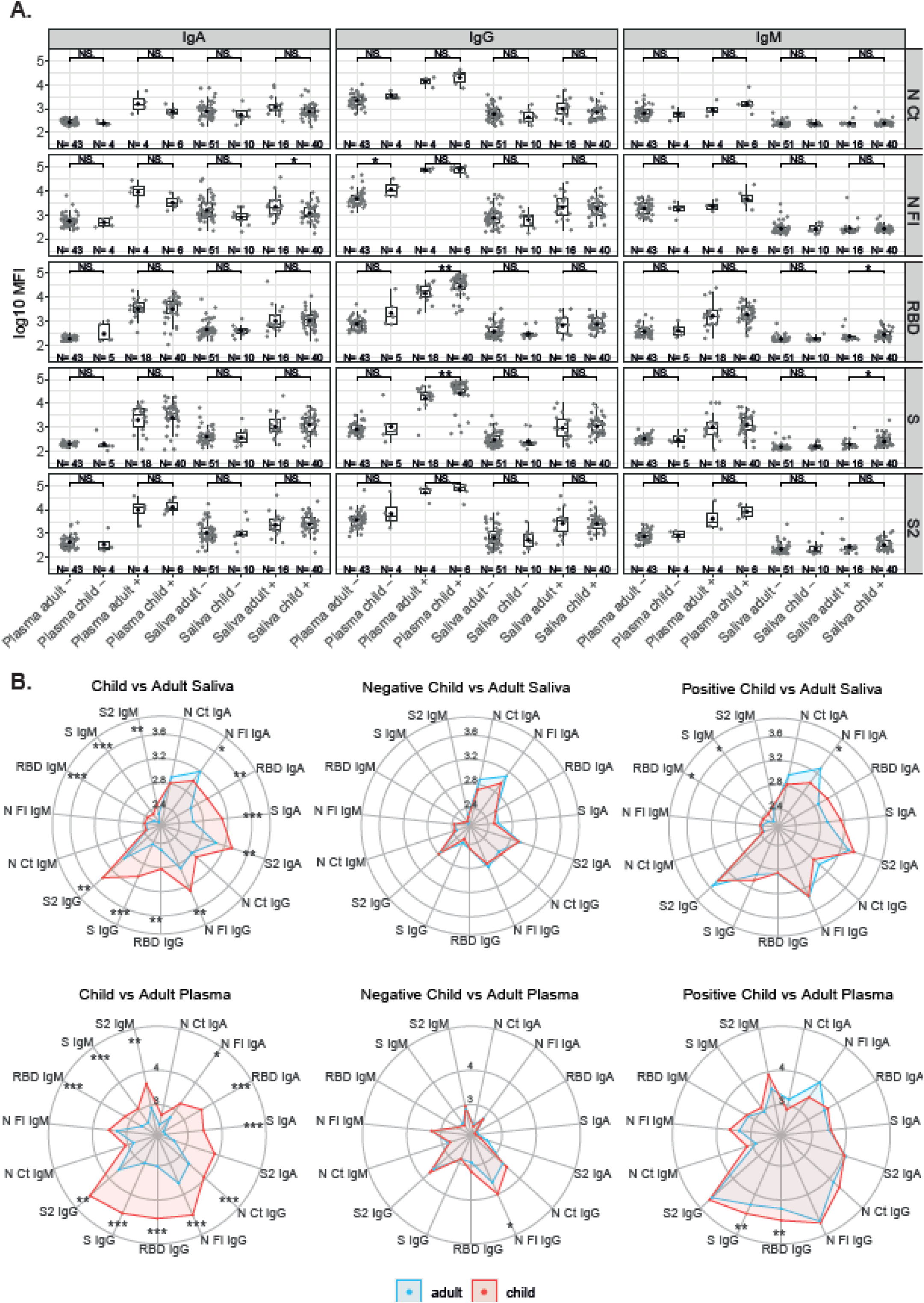
Antibody levels according to SARS-CoV-2 RT-PCR status and by age. **A**. Boxplot with saliva samples tested heat inactivated and at 1/10 dilution, and with serum (from plasma samples or dry blood spots) at 1/500. **B**. Radar charts representing the medians of antibody levels (in log_10_MFI) between child and adult plasma and saliva samples Adults are represented in blue and children in red. Median log_10_MFI antibody levels were compared by Mann-Whitney U test. Statistically significant raw p-values are highlighted with an asterisk. *** p<0.001, ** p<0.01, * p<0.05.

### Antibody levels according to presence/absence of symptoms

Among RT-PCR positive individuals, IgG levels to RBD and S were significantly higher in serum/plasma from asymptomatic than symptomatic subjects (**Figure 3A**). In general, IgG and IgA but not IgM tended to be lower in saliva and serum/plasma from SARS-CoV-2 infected subjects who developed symptoms (**Figure 3B**). Stratifying by time since symptoms onset, serum IgA and IgG to RBD and IgG to S were higher 14 days after onset of symptoms. In contrast, IgM to RBD, S and S2 in saliva were lower 14 days after onset of symptoms (**Figure 3C**).

**Figure 3.**
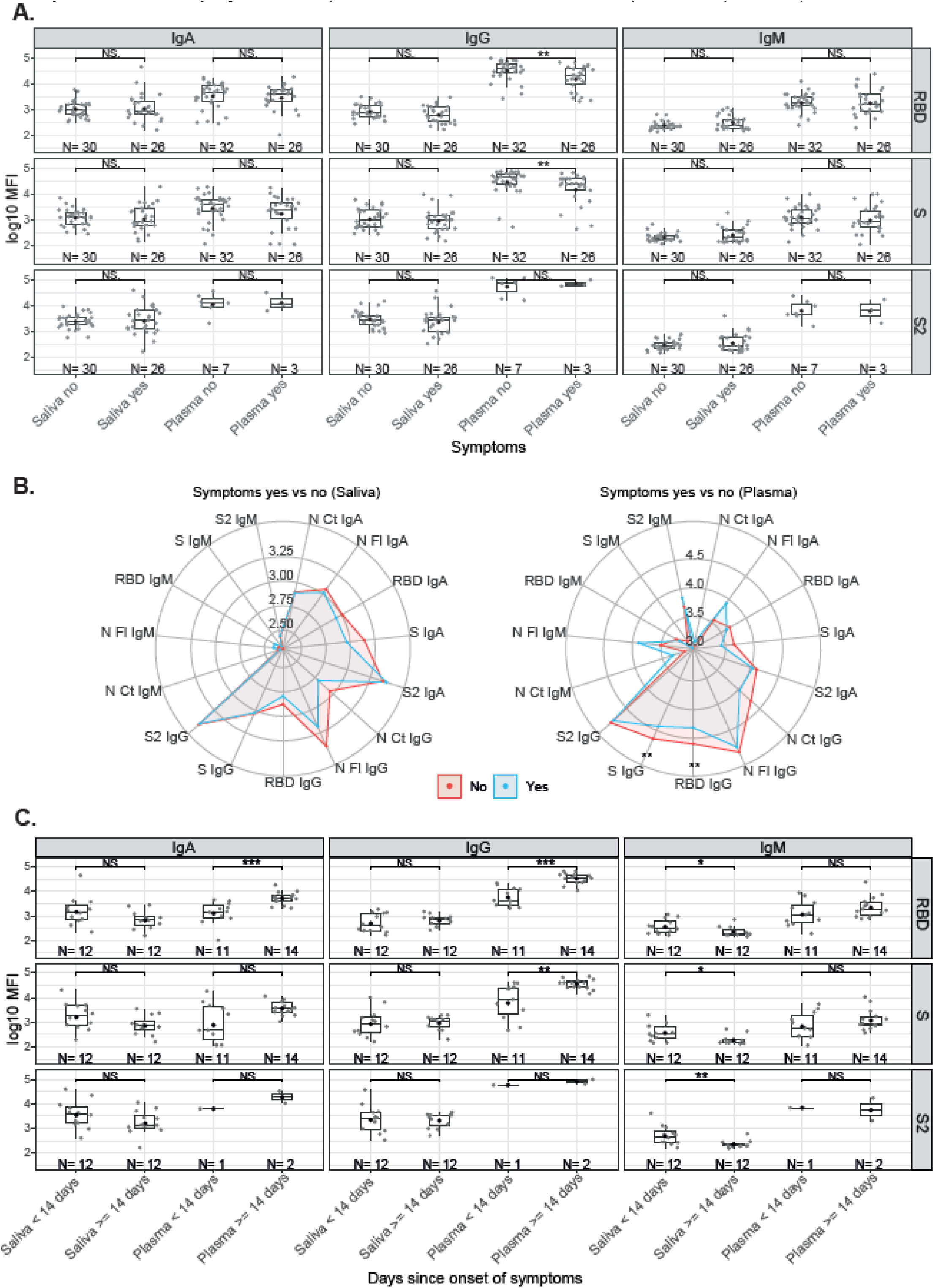
Antibody levels by symptoms in SARS-CoV-2 positive individuals. **A**. Boxplot with saliva samples tested heat inactivated and at 1/10 dilution, and serum (from plasma samples or dry blood spots) at 1/500. **B**. Radar charts representing the medians of antibody levels (in log_10_MFI) between symptomatic (blue) and asymptomatic (red) in serum/plasma and saliva. **C**. Time since onset of symptoms. Median log_10_MFI antibody levels were compared by Mann-Whitney U test. Statistically significant raw p-values indicated with an asterisk. *** p<0.001, ** p<0.01, * p<0.05.

### Correlation of antibody levels between saliva and serum/plasma samples

The pattern of antibody responses in serum/plasma versus saliva samples varied depending on the Ig isotype and antigen (**Figure 4A**). Relative antibody levels in RT-PCR positive individuals were higher in serum/plasma than saliva samples, but in RT-PCR negatives IgA levels were higher in saliva than serum/plasma samples. There was a statistically significant correlation between serum/plasma and saliva levels for all antibody isotypes against S antigens and also for IgG and IgM to N FL (**Figure 4B**). The strongest correlations were for IgG and IgA to S, followed by IgG to S2, and IgA and IgM to RBD.

**Figure 4.**
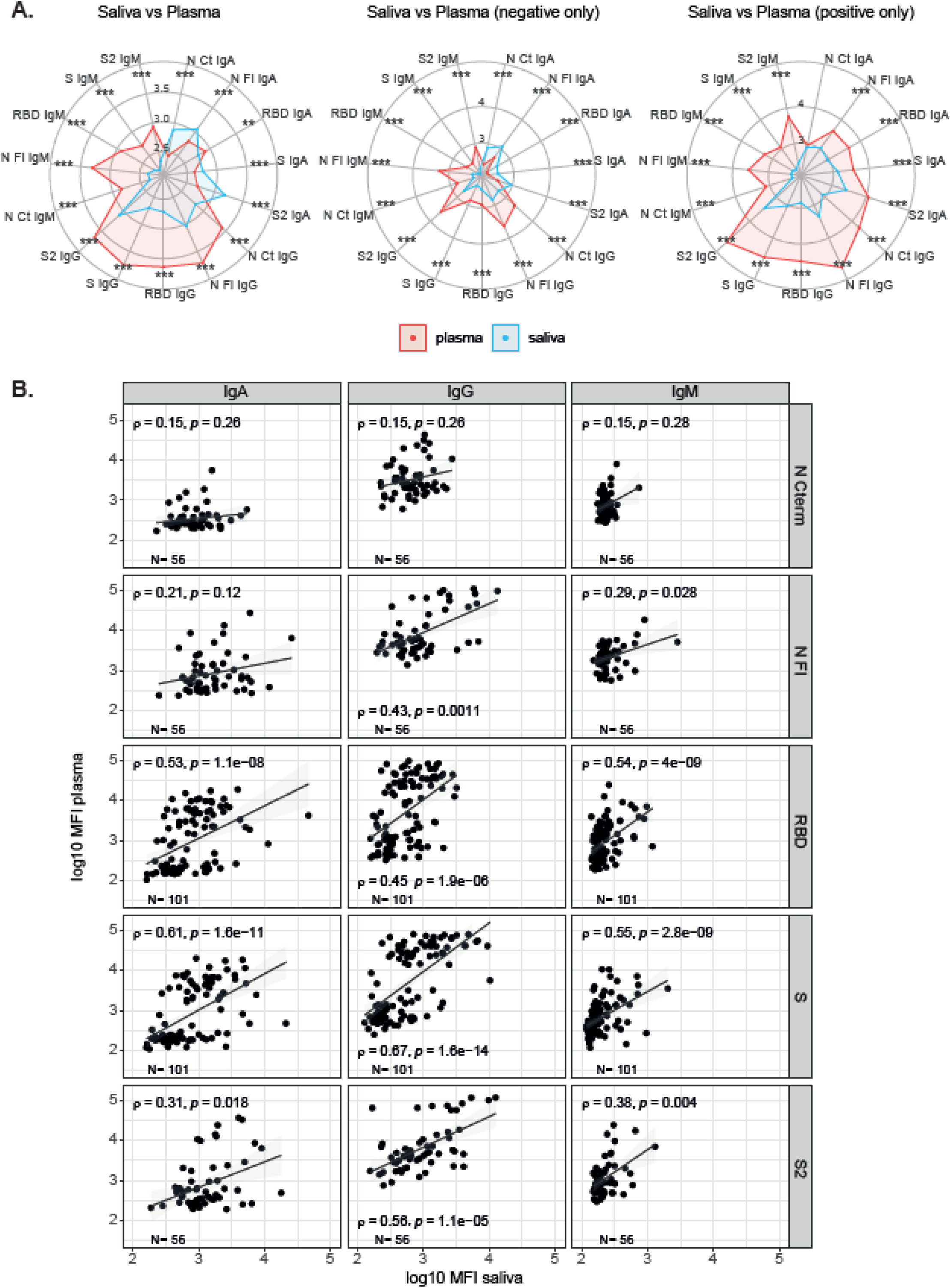
Comparison of antibody levels in serum/plasma and saliva samples. **A**. Radar charts representing the medians of antibody levels (in log_10_MFI) in serum/plasma (red) and saliva (blue). **B**. Correlations differ per Ig/Ag. X axis shows the saliva levels at 1/10 dilution, inactivated; Y axis show serum/plasma levels at 1/500 dilution, not inactivated. Median log_10_MFI antibody levels were compared by Mann-Whitney U test. Statistically significant raw p-values are highlighted with an asterisk. *** p<0.001, ** p<0.01, * p<0.05.

### Multimarker analysis of antibody responses

Combining all the antibody isotype and antigen responses per sample (**Figure 5A**) and per individual (**Figure 5B**), and considering RT-PCR status, age and symptoms, a hierarchical clustering heatmap analysis revealed different patterns. Most samples from RT-PCR positive individuals clustered together (**Figure 5A & B**) and clusters by type of samples (saliva or serum/plasma) were also observed (**Figure 5A**). RT-PCR positive individuals tended to have the wider breadth of high-level antibody responses (right side) particularly intense anti-S and anti-RBD responses in serum/plasma samples, but clusters of high responses also mapped with RT-PCR negative individuals, including some N FL, N CT and S2 IgG and IgM serum/plasma (center Fig 5A) or IgA saliva (left Fig 5A) responders. Intensity of responses was generally lower for IgM particularly in saliva, which could be influenced by the inactivation. No clear clustering was observed according to symptoms, while higher responses appeared to predominate more in children than adults.

**Figure 5.**
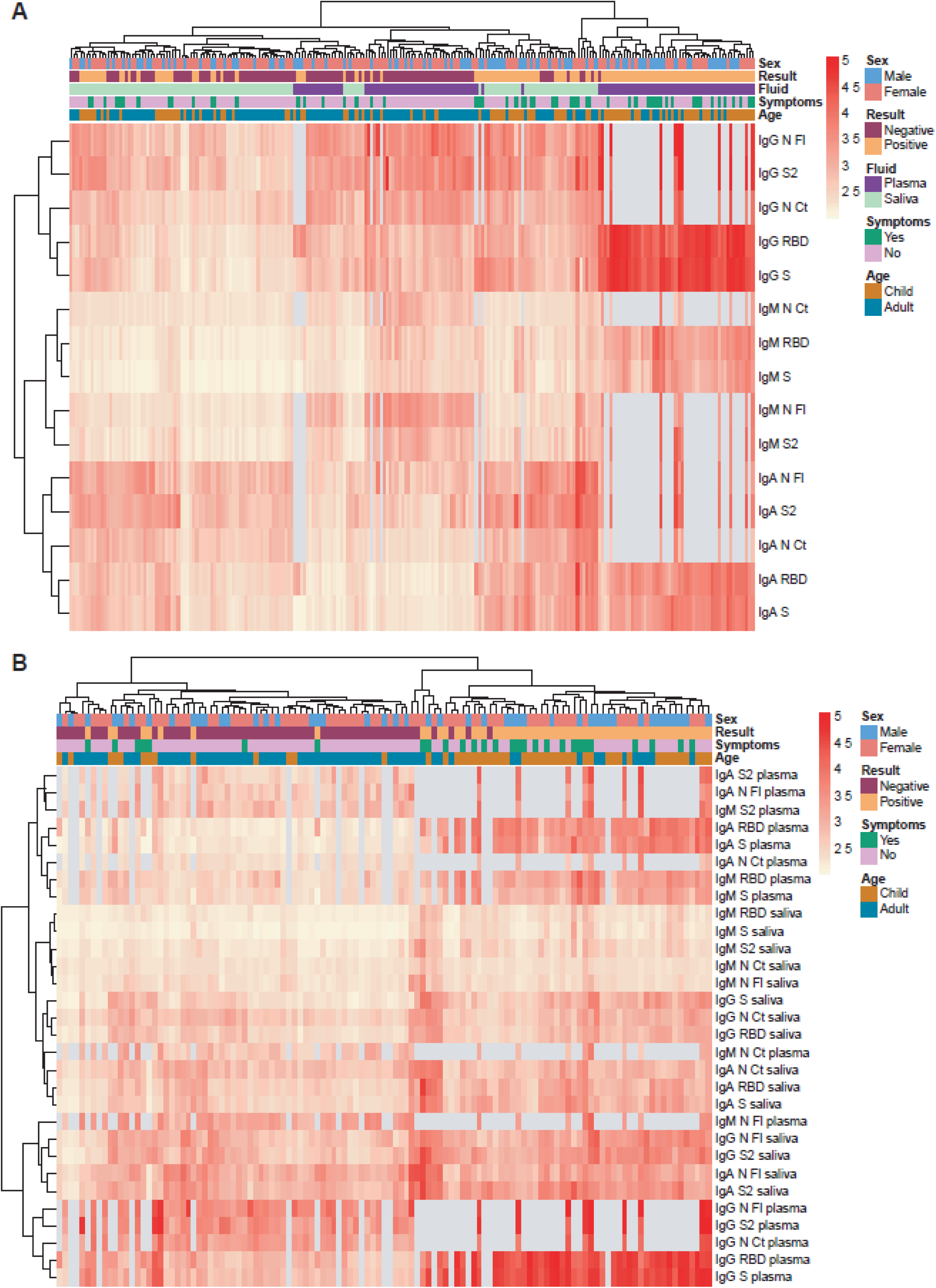
Heatmap with hierarchical clustering. Saliva samples were tested heat inactivated and at 1/10 dilution, and serum (from plasma samples or dry blood spots) at 1/500. **A**. Per sample (each column) **B**. Per individual (each column). Light grey represents missing data, as more samples were tested for S and RBD than for the rest of antigens.

## DISCUSSION

We showed that significantly higher levels of saliva antibodies to SARS-CoV-2 could be measured from RT-PCR-confirmed cases than from RT-PCR-negative individuals with our high-throughput multiplex qSAT assays. Most antibody responses correlated significantly between saliva and serum/plasma samples but Spearman coefficients were moderate/moderate-high (rho<0.7) and depended on the Ig isotype and antigen. These variable correlations would indicate different dynamics of antibody responses in blood versus mucosal tissues^8,15,16^. This suggests that responses that may not be detected in serum/plasma could be patent in saliva, and vice versa.

This makes the saliva antibody assay relevant and complementary to serology assays. The fact that saliva antibody levels were much lower but that it is a non-invasive and easy to use approach in the field compared to nasal swabs or blood pricking, represents a trade off between accuracy versus quicker and wider deployability that makes it valuable for pediatric studies.

We examined potential reasons for less discrimination between infected and non-infected subjects in saliva than in serum/plasma samples On one hand, low antibody levels in some infected individuals could be due to non-responsiveness^17^ or because a too recent exposure, as seen by higher IgGs two weeks after RT-PCR diagnosis. Low levels in saliva may not be explained by inappropriate sample collection as we used the Oracol devices that yield higher titres of total antibodies compared with other saliva/oral fluid sampling methods^18^ and are well accepted across age groups^19^. However, the quantity of antibody levels measured could have been affected by heat inactivation^8^, therefore other methods based on Triton X-100 incubation would be preferred. On the other hand, medium-high Ig levels in saliva samples from RT-PCR negative and/or serum/plasma seronegative individuals could be indicative of a previous or current exposure to SARS-CoV-2 that was not detected by RT-PCR or by serology (done by less sensitive RDT methods), i.e. false negatives. Indeed, being a study that recruited contacts of RT-PCR positive cases, it is not unreasonable that saliva serology could be more sensitive to detect infections with low or fast-resolving viral loads that might only induce a local mucosal response able to control viremia without the need to elicit a systemic response^20^. In those cases, SARS-CoV-2-specific serum IgA titers may last shorter whereas serum IgG titers might remain negative or become positive later after symptom onset while mucosal IgA might be more patent. Thus, in addition to serum IgA and IgG, measurement of SARS-CoV-2-specific saliva IgA should be considered to better estimate the percentage of individuals who have experienced coronavirus infection^21^. Alternatively, the detection of SARS-CoV-2 antibodies in saliva of RT-PCR negative people could relate to crossreactivity to common cold HCoV that are more common in children than in adults. Higher levels of pre-existing IgG to HCoV have been proposed as one potential explanation for the lower COVID-19 incidence in children^2,3^. We have seen that antibodies to N FL, followed by S2, are more present in pre-pandemic samples and be more cross-reactive^13^. These antibodies may also be more prevalent in saliva than in serum/plasma.

We investigated other factors that could be associated with the antibody responses in saliva. We observed different patterns to what is reported in cross-sectional population studies, where adults and symptomatic individuals tend to have higher antibody levels than children and asymptomatic ones, respectively. This could be related to the population under study, who are mostly children infected cases and their contacts. Higher levels of anti-S IgG among asymptomatic individuals could indicate protection against disease in infected individuals, and higher IgG and IgM levels in children could also be related to a stronger immunity. The levels of antibodies in relation to days after positive RT-PCR or days since symptoms onset reflect the kinetics whereby saliva IgM are the first to appear and decay, while serum IgA and IgG increase later. Lower viral load has been associated with faster antibody kinetics^22^. In relation to sex, the higher levels of saliva IgG to N and RBD in RT-PCR positive male children than female children could reflect sex-related differences in viral load (marker of exposure) or the ability to induce better immunity (marker of protection). This pattern is in contrast with serological studies in adults that did not find differences in males and females with mild or no symptoms^23^, and could again reflect disparate dynamics of mucosal versus systemic responses, potentially affected by sex.

The significant role of mucosal immunity and, particularly, of secretory and circulating IgA antibodies in COVID-19, is becoming more apparent, and could be exploited for beneficial diagnostic, therapeutic, or prophylactic purposes (vaccines)^7^. This supports the importance for screening antibodies in saliva in addition to serum. There is increasing evidence in favor of a key role for IgA in early virus neutralization^24^: (i) early SARS-CoV-2-specific responses are typically dominated by the IgA isotype, (ii) peripheral expansion of IgA-plasmablasts with mucosal-homing shortly after the onset of symptoms and peak during the third week of the disease, and (iii) IgA may contribute to a much larger extent to virus neutralization as compared to IgG^16,25^.

A study limitation was that seropositivity thresholds could not be estimated with pre-pandemic saliva samples due to lack of access to them, and thus sensitivity and specificity could not be established for the saliva assays by standard methods. Using RT-PCR negative pandemic samples was somewhat useful for serum/plasma samples, but with saliva there was substantial overlap between antibody levels in infected and non-infected individuals. In addition, being pandemic samples, we cannot ascertain that they were not previously exposed at low levels and therefore this approach is not optimal. This constraint could be overcome in follow up studies by assessing seroconversion in consecutive samples calculating the fold change increase in levels (e.g. ≥4) over a given study period^16^ and, in future, access to pre-pandemic samples in international biobanks will be sought. Finally, there could be some imbalance between age and infection that could affect the antibody results. However, we analyzed the effect of age stratifying by infection status to take that into account.

In conclusion, antibody levels in saliva measured with our high-throughput qSAT assay largely correlated with those in serum/plasma from individuals with confirmed RT-PCR diagnosis of SARS-CoV-2 infection, depending on the antigen. Higher antibody levels found in asymptomatic individuals and in children, particularly among males, could indicate protection against disease and stronger immunity. This non-invasive field deployable antibody screening will be useful to establish the percentage of people who have been exposed to SARS-CoV-2 in future epidemiological surveys, particularly in children. Furthermore, considering the significant correlation between saliva and serum/plasma of antibodies levels to spike, it could also be an attractive tool to monitoring induction and maintenance of vaccine responses as part of large-scale immunization campaigns.

## Data Availability

The raw data supporting the conclusions of this article will be made available by the authors upon request

## Acknowledgements

The project has been funded by Stavros Niarchos Foundation (SNF), Banco Santander and other private donors of KidsCorona. Some serum/plasma samples were tested against RBD protein produced in lentivirus by the P. Santamaria lab, IDIBAPS, and Jordi Chi contributed for production of N proteins (through PID2019-110810RB-I00 grant to L. I.). ISGlobal receives support from the Spanish Ministry of Science and Innovation through the “Centro de Excelencia Severo Ochoa 2019-2023” Program (CEX2018-000806-S), and support from the Generalitat de Catalunya through the CERCA Program. CISM is supported by the Government of Mozambique and the Spanish Agency for International Development (AECID).

## SUPPLEMENTARY FIGURES

**Figure S1.**
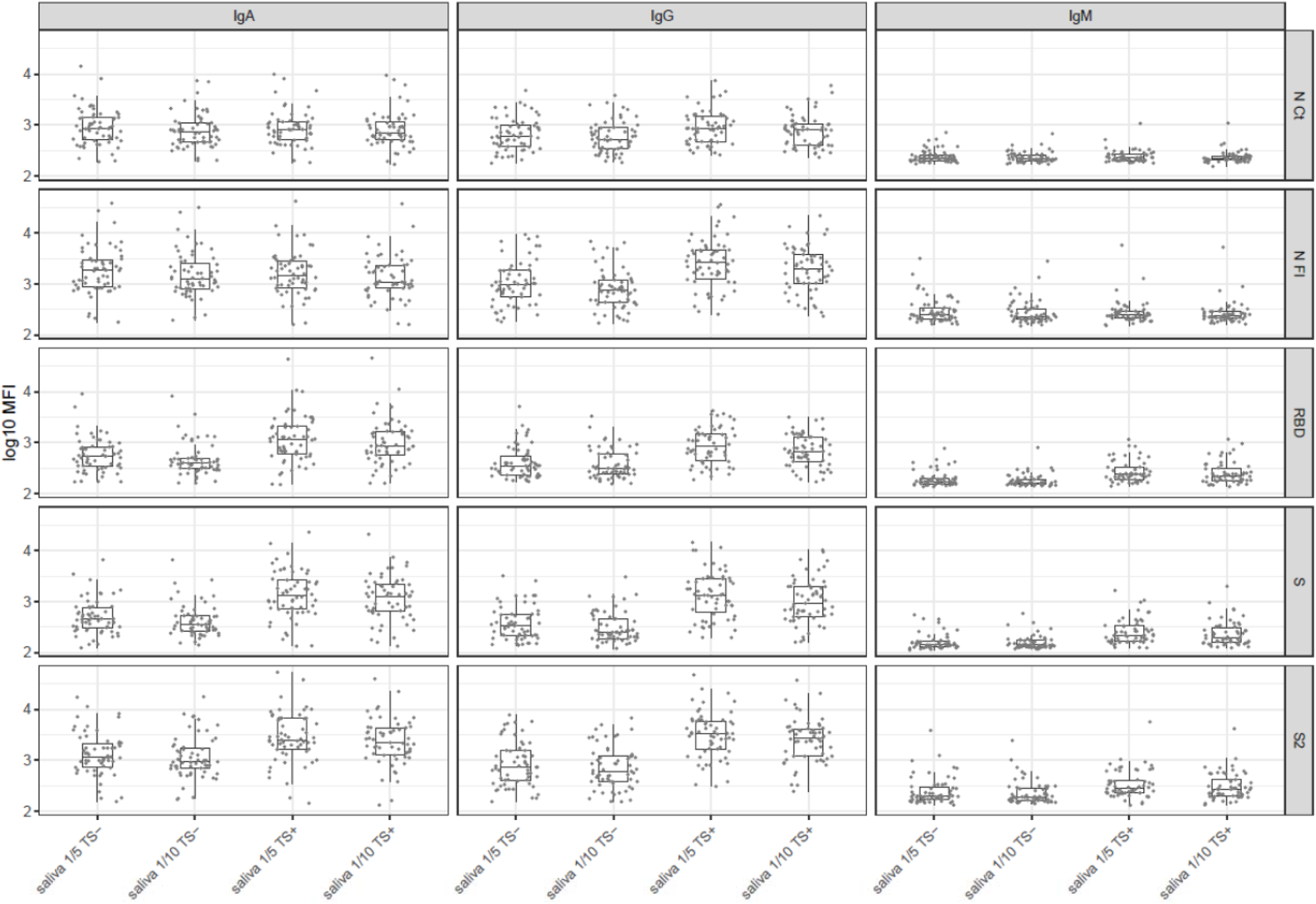
Selection of dilution of saliva 1/5 vs. 1/10 in positive and negative test samples (TS).

**Figure S2.**
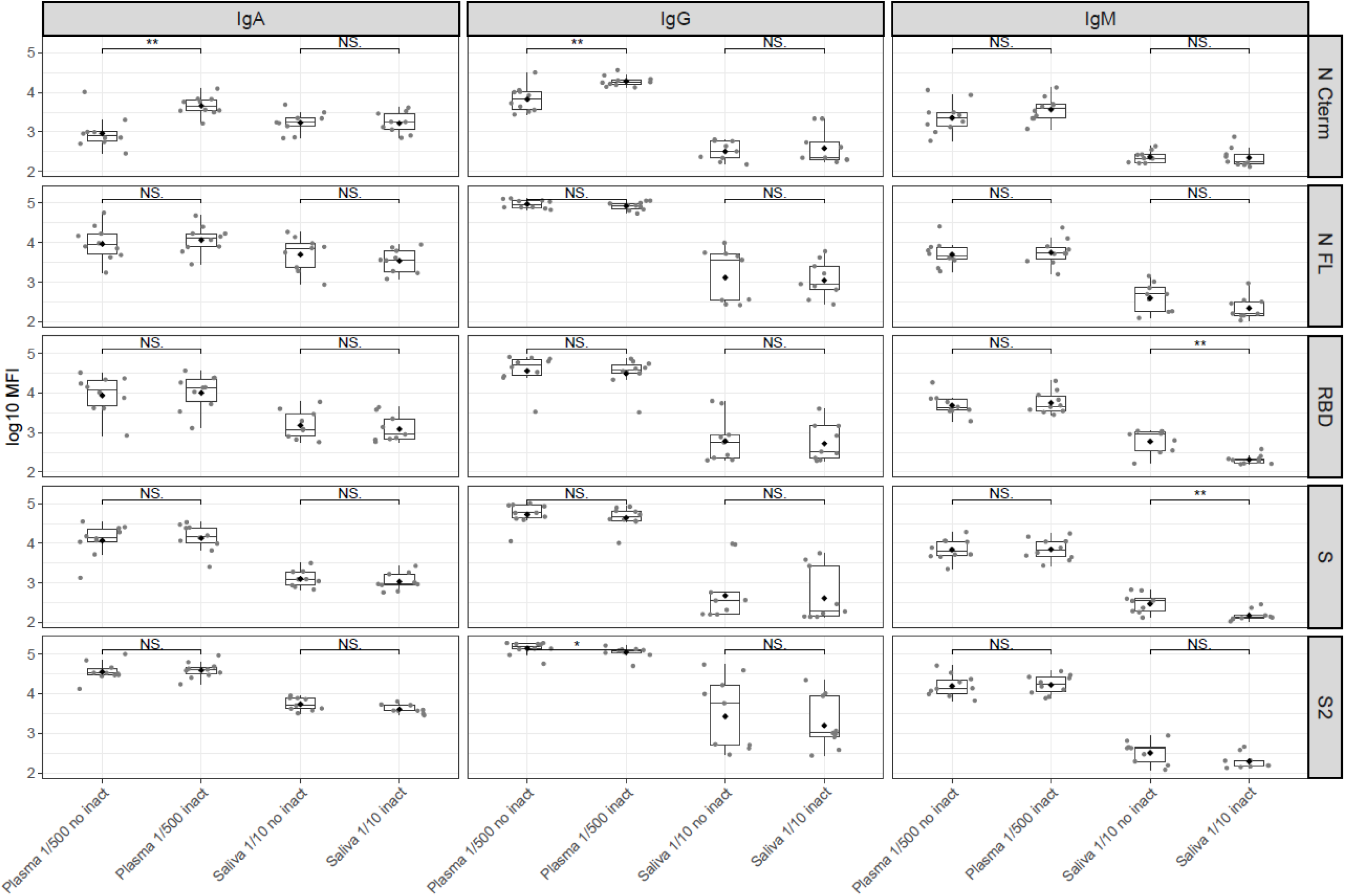
Effect of heat inactivation in saliva and serum/plasma samples.

**Figure S3.**
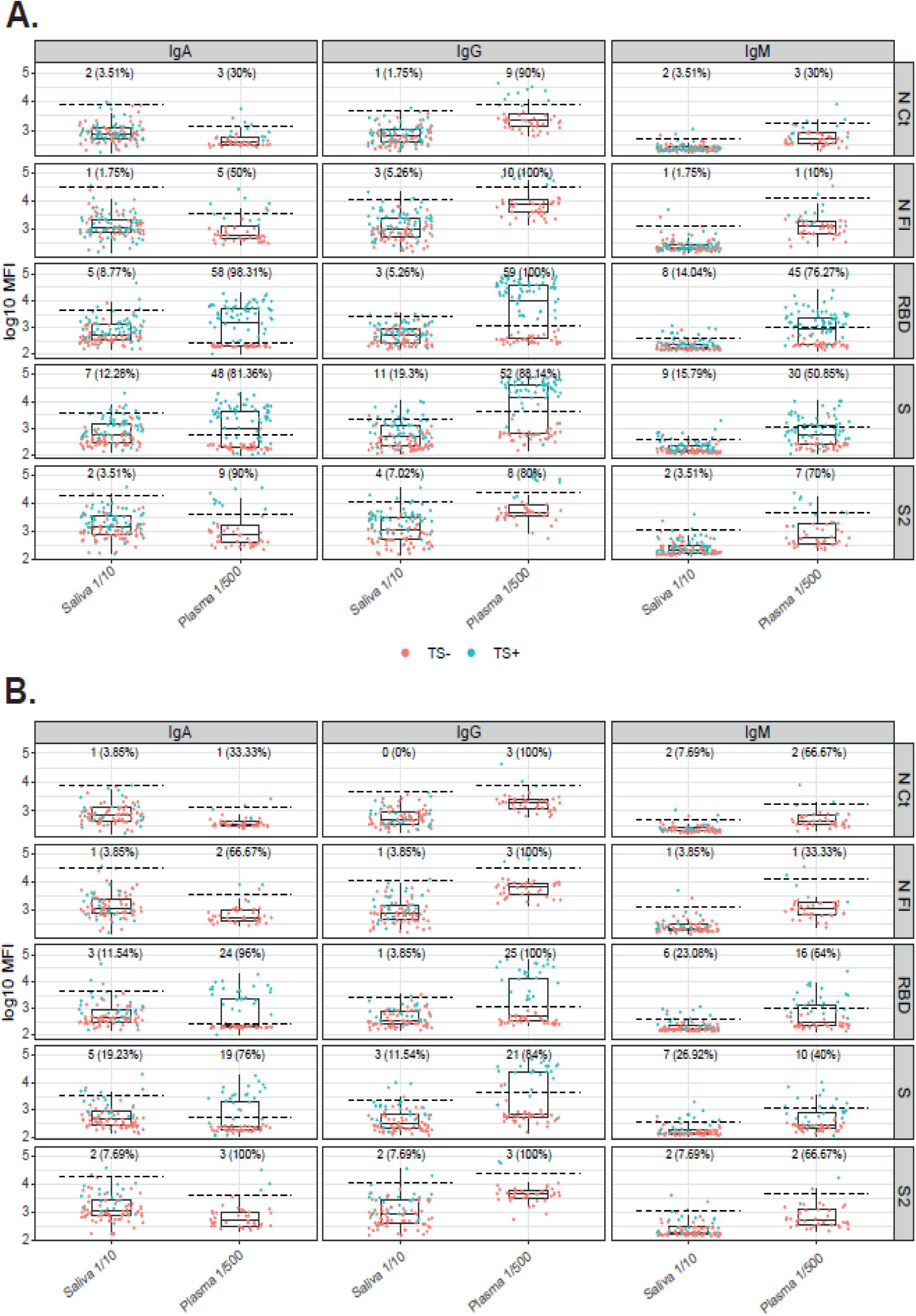
Exploring seropositivity cutoffs calculated with negative pandemic samples. Cutoffs estimated with SARS-CoV-2 RT-PCR negative pandemic samples (TS-) discriminated up to 100% the positive individuals (TS+) depending on the antigen in serum/plasma but not saliva samples due to the overlap in antibody levels **(A)**. Cutoffs estimated with the RT-PCR negative pandemic samples in symptomatic patients discriminated positive individuals in more antigens **(B)**. Salivas were tested at 1/10 dilution and serum/plasmas at 1/500 dilution.

**Figure S4.**
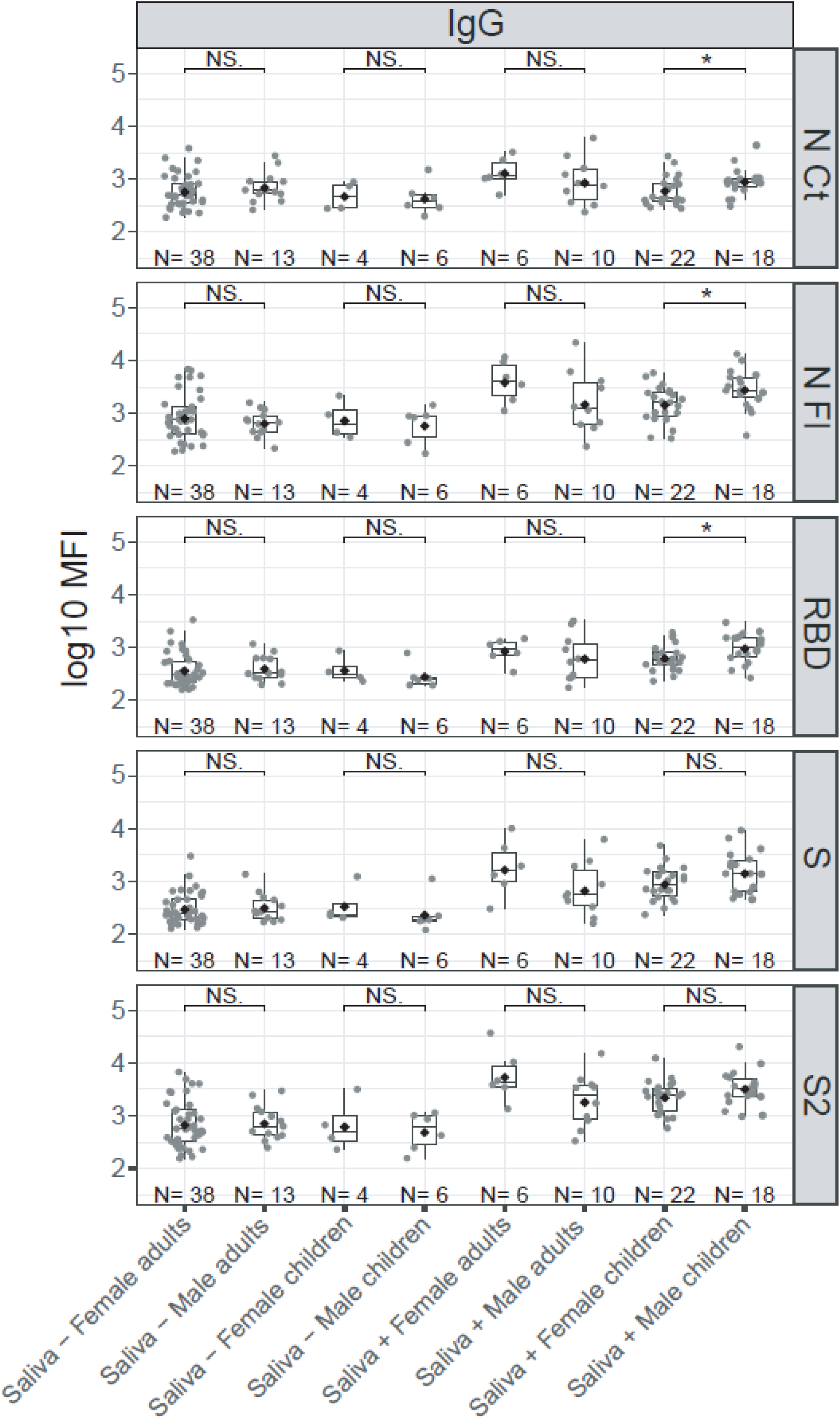
Comparison of antibody levels in saliva samples stratified by sex and age.

